# LONG-TERM OUTCOMES OF ENDOSCOPIC ULTRASOUND-GUIDED RADIOFREQUENCY ABLATION (EUS-RFA) FOR ADVANCED PANCREATIC AND PERIAMPULLARY ADENOCARCINOMA

**DOI:** 10.1101/2021.12.11.21267660

**Authors:** Nirav Thosani, Putao Cen, Julie Rowe, Sushovan Guha, Jennifer Bailey-Lundberg, Dimpal Bhakta, Prithvi Patil, Shahrooz Rashtak, Roy Tomas Davee, Srinivas Ramireddy, Curtis J. Wray

## Abstract

**Background:** Long term prognosis for pancreatic adenocarcinoma (PDAC) remains especially poor with an overall 5-year survival rate less than 9%. Endoscopic ultrasound (EUS) guided RFA (EUS-RFA) is an emerging technology and limited data exist regarding long-term outcomes of EUS-RFA for PDAC. In addition to thermal-induced coagulative necrosis and tissue damage, radiofrequency ablation (RFA) has potential to stimulate the host’s antitumor immunity. The aim of this study is to report long-term outcomes of EUS-RFA for unresectable PDAC.

**Methods:** Retrospective chart review of adult patients with an established diagnosis of locally-advanced or metastatic PDAC undergoing EUS-RFA between October 2016 to March 2018 with long term follow up (>30 months). Patients included in the review underwent a total of 1-4 RFA sessions using the Habib EUS-RFA radiofrequency catheter. All patients were concurrently undergoing standard of care chemotherapy.

**Results:** 10 patients (median age 62 years, male 70%) underwent EUS-RFA (Table 1). Location of the primary PDAC was in the head (4), neck (2), body (2), and tail (2). A total of 22 RFA sessions were performed with a range of 1-4 RFA sessions per patient. RFA was technically successful in all RFA sessions (100%). There were no major adverse events (bleeding, perforation, infection, pancreatitis) in immediate (up to 72 hours) and short-term follow up (4 week). Mild worsening of existing abdominal pain was noted during post-procedure observation in 12/22 (55%) of RFA treatments. Follow-up imaging after RFA treatment was available in 8/10 patients. Tumor progression was noted in 2 patients, whereas tumor regression was noted in 6 patients (>50% reduction in size in 3 patients). Median survival for the cohort was 20.5 months (95% CI, 9.93 to 42.2 months). Currently, 2 patients remain alive at 53 and 73 months follow-up since initial diagnosis. One patient had 3 cm PDAC with encasement of the portal confluence, abutment of the celiac axis, common hepatic and superior mesenteric artery. This patient had significant reduction in tumor size and underwent standard pancreaticoduodenectomy.

**Conclusion:** In our experience, EUS-RFA was safe, well-tolerated and could be concurrently performed with standard of care chemotherapy. In this select cohort, median survival (20.5 months) was improved when compared to published survival based upon SEER database and clinical trials. Future prospective trials are needed to understand the role of EUS-RFA in overall management of PDAC.

## INTRODUCTION

### Background

Pancreatic ductal adenocarcinoma (PDAC) remains a leading cause of cancer death in the United States and is highly resistant to therapy [1, 2]. Surgical resection remains the only curative option, but more than 80% of patients present with unresectable disease highlighting the urgent need for improved neoadjuvant therapeutic options [3]. Unfortunately, even among those who are candidates and undergo surgical resection, the reported median survival is 15-23 months, with a 5-year survival less than 20% [4–6]. These dismal survival rates over the past several decades remain disappointing. Despite improvements in diagnostic imaging, surgical technique and chemotherapeutic options, only modest improvements in PDAC survival have been reported. It remains clear that surgical resection is a prerequisite for achieving long-term survival; thus, innovative treatments to increase odds for R0 resection are needed to improve overall survival in PDAC [7].

Currently patients diagnosed with locally-advanced, unresectable or metastatic PDAC are only candidates for systemic chemotherapy and/or chemoradiation. As defined by the National Comprehensive Cancer Network (NCCN), locally-advanced pancreatic cancer (LAPC) criteria include: (1) > 180 degrees contact of superior mesenteric artery or common hepatic artery, (2) aortic involvement or (3) unreconstructable superior mesenteric vein/portal vein due to tumor involvement or occlusion [8]. In highly selected patients with locally-advanced disease, neoadjuvant chemotherapy (NAC) and chemoradiation has resulted in diagnostic downstaging resulting in surgical resection in 30-50% of cases [9, 10] and progression in 20% of cases. For patients that progressed after NAC, the median survival is < 18 months. These statistics highlight the incredible potential for PDAC to adapt and resist NAC in a subset of patients. In those eligible for surgical resection, NAC is highly recommended by the NCCN as probable benefits include tumor stage downsizing and increased probability of a margin-negative resection [11–13].

At present, conventional treatment modalities for PDAC are limited to chemotherapy, radiation and surgery; these treatments have been used for the last 20+ years [14]. Recent advances in endoscopic therapy may provide an additional therapeutic modality. Several percutaneous and endoscopic ultrasound (EUS) guided ablative techniques have been explored for the treatment of pancreatic lesions such as alcohol injection, photodynamic therapy, and laser ablation [15]. These techniques are less-invasive which enhances feasability and may be safer in patients who are poor candidates for surgical resection. EUS guided radiofrequency ablation (EUS-RFA) is one of the newer techniques currently available [16]. EUS-RFA application consists of alternating current with a frequency of 350–500 kHz (coincidentally the frequency range of radio broadcasts as well) to the target tissue via a special electrode located at the tip of the endoscope. Alternating current causes vibratory movement of ionic particles in the abutting and adjoining tissue resulting in generation of heat. RFA induces not only local disruption of the tumor by heat, but also produces intratumoral localized coagulation necrosis which results in the release of large amounts of cellular debris [17]. Local and systemic release of cellular debris is postulated to be a tumor antigen source that can trigger a host adaptive immune response targeting PDAC. The release of pathogenic “noxa” into the body induces a strong inflammatory response with elevated levels of IL-6, HGF, and VEGF [18]. RFA has recently been recognized for its potential in palliative treatment of malignant biliary strictures [19, 20]. Based on published data, RFA provides palliation and may increase survival [21, 22]. These data suggest RFA may be considered as an additional neoadjuvant therapy for locally advanced pancreatic cancer as a novel method to combine local and systemic immune-modulatory effects. Our hypothesis is that EUS-RFA is a safe treatment modality and may improve survival for unresectable or locally-advanced pancreatic ductal adenocarcinoma.

## METHODS

We evaluated adult patients with an established, histologically confirmed diagnosis of unresectable or locally-advanced PDAC. Inclusion criteria were: (1) diagnosis of PDAC, (2) locally-advanced PDAC, (3) contraindication or refusal of radiation therapy and (4) patients with stable advanced disease (defined as greater than 6 months chemotherapy). Eligible patients underwent EUS-RFA at the University of Texas Health Sciences Center at Houston (UTHealth) between October 2016 to March 2018 with long term follow up (>30 months). All patients were enrolled in a prospective EUS-RFA study (HSC-MS-18-0192) approved by the UTHealth Institutional Review Board.

All patients were concurrently undergoing standard of care systemic chemotherapy, either modified FOLFIRINOX: Oxaliplatin 85 mg/m2 IV over 2 hours, Leucovorin 400 mg/m2 IV over 2 hours, Irinotecan 150 mg/m2 IV over 90 minutes, Fluorouracil 2,400 mg/m2 IV continuous infusion (CI) over 46 hours beginning on Day 1 and repeat this cycle every 14 days or Gemcitabine/nab-Pacliataxel: Gemcitabine 500-1000mg/m2 over 30minutes Nab-Paclitaxel 125mg/m2 over 30minutes +/- Cisplatin 25mg/m2 over 1hour with 1liter NS hydration. Weekly, 3 weeks on and one week off (Day 1,8, 15 every 28 days).

Patients in this study underwent upper endoscopy using therapeutic Pentax or Olympus scope. The EUS-RFA procedure was performed using the Habib ablation catheter. Prior to RFA, a standard fine needle aspiration (FNA) was performed. After the FNA, a 19- or 22-gauge probe was advanced into the target lesion. Using ultrasound guidance, transduodenal or transgastric electrode needle probe placement into the target lesion was performed. Color Doppler scanning was performed in order to avoid nearby or adjacent blood vessels. The RFA parameters included treatment of target lesion (10-15 W) to a measured electrical resistance of 200 Ohm or a rapid change in Ohms. Treatment was confirmed by the gastroenterologist who noted either liquefaction or bubbling noted with real-time EUS. Patients received up to a total of 4 EUS-RFA sessions/treatments (1 session every 2-3 weeks) based upon the completeness of ablation as determined by the interventional gastroenterologist.

The primary endpoint of this study was technical success of the EUS-RFA procedure. Secondary endpoints included: (1) safety, side-effects, adverse events and (2) overall survival. The date of PDAC diagnosis and date of last contact or death were used for Kaplan Meier analysis to measure survival (STATA 16, College Station, TX). The Response Evaluation Criteria in Solid Tumors (RECIST) was used to evaluate radiographic treatment [23].

## RESULTS

In this prospective study, a total of 10 patients (mean age 62 years) underwent EUS-RFA (**Table 1**). The majority of patients in this study were male (70%) and Caucasian (80%). At the time of RFA, a majority of the patients were AJCC stage III (70%). Based upon accepted definitions of PDAC tumor anatomy, 7 patients were locally-advanced and 3 patients were metastatic. In most cases, EUS-RFA was performed as an outpatient procedure. Location of the primary PDAC was in the head (4), neck (2), body (2), and tail (2)(**Table 2**). At diagnosis, mean CA19-9 was 2004.3 (SD 2736.8) and was elevated in 7 patients (70%). Systemic chemotherapy was used in all patients (mFOLFIRINOX = 2, gemcitabine/Abraxane = 1, both mFOLFIRINOX and gemcitabine/Abraxane = 6, mFOLFIRINOX and gemcitabine/Abraxane + Cisplatin = 1). Three patients also received external beam radiation in addition to EUS-RFA.

**Table 1:** Patient demographics.

| **Variable** | **Mean** | **N** | **%** |
| --- | --- | --- | --- |
| **Age (SD)** | 62.3 (5.74) |  |  |
| **Gender** |  |  |  |
| Male |  | 7 | 0.7 |
| Female |  | 3 | 0.3 |
| **Race** |  |  |  |
| Caucasian |  | 8 | 0.8 |
| Asian/Other |  | 2 | 0.2 |
| **Stage** |  |  |  |
| III |  | 7 | 0.7 |
| IV |  | 3 | 0.3 |
| **Anatomy** |  |  |  |
| LAPC |  | 7 | 0.7 |
| Metastatic |  | 3 | 0.3 |
| **EUS Setting** |  |  |  |
| Inpatient |  | 1 | 0.1 |
| Outpatient |  | 9 | 0.9 |
| **RFA Duration** |  |  |  |
| Time (SD) | 32.6 (14.5) |  |  |

**Table 2:** Characteristics of pancreatic lesions, number of RFA sessions and overall survival in patients undergoing EUS-RFA (include survival from 1^st^ RFA)

| Patient | Size of Mass at Diagnosis (mm) | Location | Number of RFA Sessions | Deceased | Survival (months) |
| --- | --- | --- | --- | --- | --- |
| 1 | 40 x 33 | Head | 2 | Yes | 9 |
| 2 | 35 x 35 | Neck | 3 | Yes | 9 |
| 3 | 30 x 20 | Head | 1 | Yes | 10 |
| 4 | 44 x 40 | Body | 4 | Yes | 16 |
| 5 | 53 x 39 | Tail | 1 | Yes | 17 |
| 6 | 59 x 47 | Neck | 1 | Yes | 23 |
| 7 | 68 x 50 | Body | 1 | Yes | 27 |
| 8 | 28 x 25 | Head | 3 | No | 48 |
| 9 | 21 x 18 | Tail | 4 | Yes | 42 |
| 10 | 14 x 12 | Head | 2 | No | 68 |

A total of 22 EUS-RFA sessions were performed with a range of 1-4 RFA sessions per patient. There were no aborted endoscopic procedures and all 10 patients were successfully treated with EUS-RFA (100%). Mean time for the EUS procedure was 32.6 minutes (range 18-66). There were no major adverse events (bleeding, perforation, infection, pancreatitis) in the immediate (up to 72 hours) and short-term follow up (4 weeks) period. Mild worsening of existing abdominal pain was noted during post-procedure observation in 12/22 (55%) of RFA treatments. This was managed by short-term opioid therapy and extended post-procedure observation (1-4 hours) in the recovery unit. None of the patients required hospital admission following EUS-RFA. Serum CA19-9 was measured in longitudinal fashion 1 week prior to and following EUS-RFA. In the 7 patients with elevated baseline CA19-9, 12 of the 15 treatments resulted in a decrease of the subsequent measured value (**Figure 1**). This effect is most prominent in patient #5 (red line) and patient #8 (green line).

**Figure 1.**
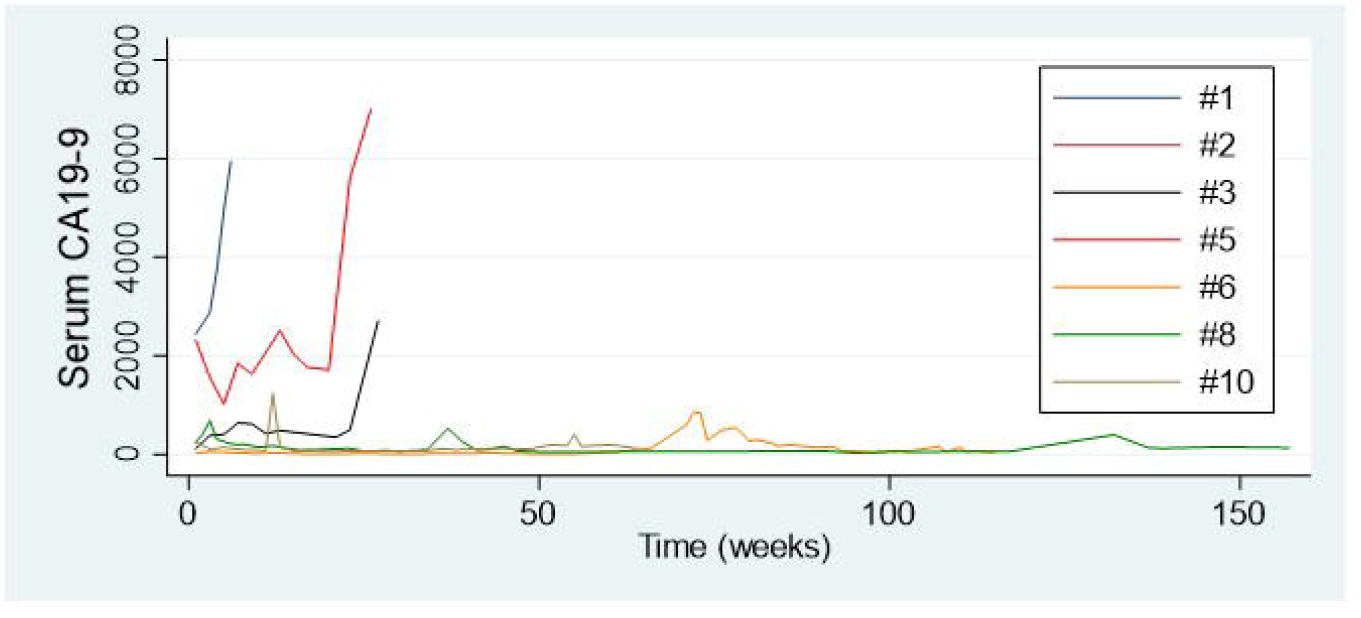
Serum CA19-9 measured following EUS-RFA treatment

Follow-up imaging after RFA treatment was available in 9 of the 10 patients. Primary tumor progression was noted in 2 patients, whereas tumor regression was noted in 7 patients (>50% reduction in size in 3 patients). Kaplan-Meier survival analysis demonstrated median survival duration of 20.5 months (95% CI: 9.93 - 42.2) (**Figure 2**). The median survival post-RFA was 13.4 months (95% CI: 0.9 – 27.8). Currently, 2 patients are still alive at 53 months and 73 months follow-up since initial cancer diagnosis. One of these patients had a 3 cm primary tumor with abutment of the portal confluence, abutment of the celiac axis, common hepatic artery and superior mesenteric artery. This patient was treated with extended systemic chemotherapy (mFOLFIRINOX 19 cycles), intensity modulated radiation therapy (50.4 Gy) and 3 EUS-RFA sessions. The tumor decreased in size with regression along the previously involved mesenteric vessels (**Figure 3**). This patient tolerated all pre-operative treatment with minimal toxicity and subsequently underwent a margin negative (R0) pancreaticoduodenectomy (alive at 53 months). Another patient was initially thought to have metastatic PDAC with a histologically proven positive mediastinal node (Stage IV). This patient underwent extended chemotherapy (3 cycles mFOLFIRINOX, 32 cycles Gemcitabine/nab-Paclitaxel and 3 cycles of Gemcitabine/nab-Paclitaxel/Cisplantin) and 2 EUS-RFA treatments. Robotic-assisted laparoscopic pancreaticoduodenectomy was performed and the surgical pathology demonstrated an ampullary adenocarcinoma. This patient remains alive with no evidence of disease at 73 months from initial diagnosis.

**Figure 2:**
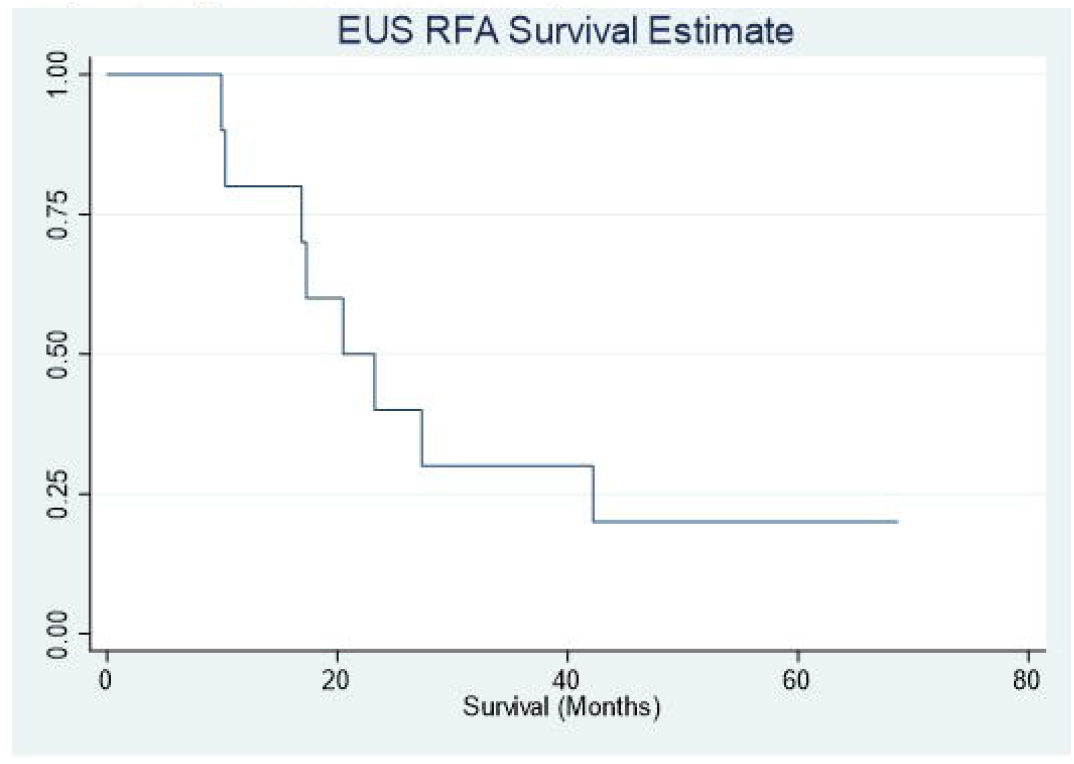
Kaplan-Meier Survival Analysis.

**Figure 3.**
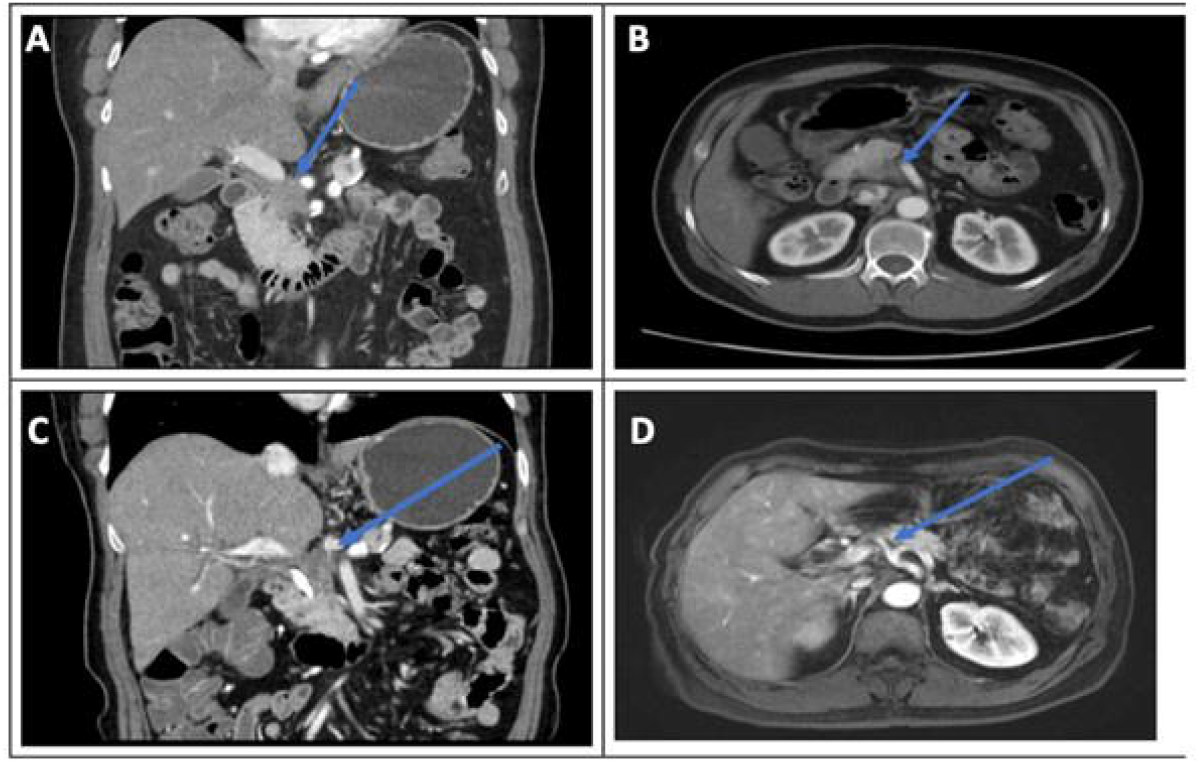
Representative computed tomography shows reduced PDAC lesion size post RFA (A) Computed tomography (CT) with pancreas head mass and evidence of portal confluence encasement. (B) CT with pancreas head mass and evidence of superior mesenteric artery abutment. (C) CT post RFA with resolution of previously seen encasement of celiac axis as evidenced by a fat plane between tumor and celiac vessels. (D) Magnetic resonance imaging post RFA of PDAC with no further evidence of tumoral abutment of celiac axis, superior mesenteric artery and superior mesenteric vein.

## CONCLUSIONS

In our experience, EUS-RFA was safe, well-tolerated and concurrently performed with standard of care PDAC chemotherapy treatment. There were no significant side effects or adverse events noted in this small series. The observed overall survival (20.5 months) in this cohort is encouraging, despite advanced stage disease (III or IV) in all patients. This appears to be an improvement to published expected survival (9-12 months) for LAPC treated with chemotherapy alone [24]. These results are also comparable to locally-advanced patients treated with radiation dose escalation after induction chemotherapy (median overall survival 17.8 months) [25]. However, escalated radiation dose protocols resulted in higher rates of known side effects such as abdominal pain, nausea, vomiting, diarrhea or fatigue. In addition, escalated radiation dose results in anemia requiring transfusion in 13% of PDAC patients [30]. Thus, the feasibility and tolerance of >50.4 Gy radiation treatment is likely limited in older, debilitated LAPC patients. Although median survival may be either equivalent or possibly better when comparing our small cohort to dose-escalated radiation, EUS-RFA may be both safer and better tolerated in LAPC patients with marginal performance status.

These data are timely as the widespread use of ablative techniques is employed in the management of other gastrointestinal cancers; however, RFA treatment of the pancreas has been very limited. Ablation has not been widely used as therapy for PDAC due to potential risk of complications from the increased sensitivity of pancreatic tissue to thermal injury, concerns for pancreatitis and proximity to critical vascular and biliary structures. Prior to the newer EUS approach, other options were either percutaneous of surgical which may be an aggressive modality in patients with advanced PDAC. Albeit a small sample size, EUS-RFA was technically feasible to perform and well-tolerated in this PDAC patient cohort.

Several investigators have demonstrated the feasibility of EUS-RFA in porcine and animal models [26–28]. In animal models of pancreatic RFA, short-term results appeared safe as most rodent or porcine patients survived unharmed until euthanized per the research protocol. Translation into clinical use has been hindered by both endoscopic and technologic challenges. Open surgical or laparoscopic approaches may not be well-tolerated with limited feasibility since most surgeons are not trained in pancreatic ablation. Thus, endoscopic RFA may be the ideal approach when compared to traditional open or laparoscopic pancreatic ablation in which postoperative complication rates approach 25% [29]. Endoscopy also has the advantage of concurrent EUS visualization and ultrasound mode scanning. The EUS technique has been used in pancreatic neuroendocrine tumors with similar results (<10% complication rate) [30]. In a recent update from Barthet et al., 65% of patients that underwent EUS-RFA treatment of a neuroendocrine tumor of cystic neoplasm had complete disappearance of the index lesion [31]. The data from these studies suggest the EUS-RFA approach may be a durable alternative treatment modality for pancreatic neuroendocrine tumors (<3cm) and select cystic neoplasms.

PDAC-related mortality has not significantly declined over the past two decades, unlike other solid malignancies (lung, colon, breast). Multi-agent preoperative chemotherapy remains the current gold-standard, yet the majority of PDAC patients are not candidates for potentially curative surgery. At present, both chemotherapy and chemoradiation have been employed in borderline resectable and locally-advanced PDAC with mixed results [32]. As newer technologies emerge, local ablative treatments may play a role in the treatment of PDAC. As mentioned previously, EUS-RFA treatment in combination with standard chemotherapy may be beneficial for several reasons: (1) direct treatment of the PDAC through coagulative necrosis, (2) improved chemotherapeutic efficacy as the tumor may become more porous and allow systemic therapies to further penetrate into the tissues and (3) possibly enhanced, systemic antitumor immunity as necrotic PDAC released into the bloodstream may be recognized by the host immune system. Tumor ablation has been shown to enhance systemic immune-mediated effects [33, 34]. These systemic effects may help to explain the patient in this series with stage IV ampullary carcinoma who remains alive at 73 months from date of diagnosis. Obviously tumor biology plays a key role; however, it is possible that RFA sensitized the immune system to treat systemic disease.

There are several important limitations to discuss. This PDAC patient cohort is relatively heterogeneous with regards to tumor size, location and stage. This study was nonrandomized and the systemic treatment protocol was not equivalent in all patients. In this small cohort, there was no standardized approach to the timing of EUS-RFA in combination with chemotherapy and/or radiation. In addition, follow-up imaging was unavailable in 1 patient. Despite these obvious limitations, this is a mature data set with > 30 months follow-up. Notwithstanding the small sample size, this unique data adds to the available body of evidence and clinical experience that EUS-RFA is a safe treatment modality for PDAC.

These results highlight our institutional EUS-RFA experience in the treatment of pancreatic adenocarcinoma. This minimally-invasive procedure was well-tolerated in patients with advanced disease and significant comorbidities. In addition, two patients with advanced disease responded to the RFA treatment and were able to undergo potentially curative pancreaticoduodenectomy. At the time of resection, there was no appreciable scarring or fibrosis. This had been a significant operative concern as external beam radiation and fibrosis is known to interfere with radical surgery. In these two cases, the EUS-RFA did not appear to negatively influence the complex resection. This report of pancreatic RFA suggests this procedure may be a complementary treatment alongside standard chemotherapy. This new combination therapy may extend an effective treatment to a larger group of patients. This may provide an effective therapy to older and/or fragile patients that may not tolerate other treatments such as chemotherapy with dose-escalated radiation. This initial experience is promising and suggests future clinical trials are needed to understand the role and timing of EUS-RFA in the management of both localized and advanced pancreatic ductal adenocarcinoma. At present, our institution is currently conducting a single-arm, phase II trial (www.clinicaltrials.gov NCT04990609) of neoadjuvant chemotherapy plus EUS-RFA for resectable pancreas cancer.

## Data Availability

All data produced in the present study are available upon reasonable request to the authors

